# Mozambican Food Pyramid

**DOI:** 10.1101/2024.08.24.24311155

**Authors:** Ernesto Francisco Maborre Fotinho

## Abstract

**Introduction:** In order to support nutritionists and other food and nutrition education actors in Mozambique in improving the approaches of intervention messages to fight for chronic malnutrition and other nutritional related diseases that plague the country, this work brings the adaptation of the American food pyramid published in 1993 to the local reality.

**Methods:** The pyramid was built with the foods distributed in eight groups (Basic Foods, fruits, vegetables, legumes, milk, meats, fats and sugars) and in 5 levels. Each level was presented in minimum and medium portions to be consumed according to the established standard diets. The portions and the food equivalents tables were defined according to the caloric contribution of each food group using an automated spreadsheet of Microsoft Excel, version 19, and based on the principle of linear programming. The pyramid was based on two standard diets (1693 kcal and 2300 kcal) with four meals for both.

**Results:** The percentage distribution of macronutrients in both diets is within the recommended range: Carbohydrates (45 to 65%), lipids (20 to 35%) and proteins (10 to 35%). Special attention was given to the milk group by calculating the bioavailable calcium values.

**Discussion:** Factors such as food availability, dietary reference intake, and eating habits of the local population were elements of adaptation to the local context.

**Conclusion:** Further studies are needed to validate and implement the instrument so that it becomes an effective and practical tool.

**Key message:**

- What is already known on this topic: The tools that are currently used to promote healthy eating habits existing in parents present a gap regarding the portions of each food group that should be consumed, constituting a limitation for professionals in the area, who have been struggling to seek information on graphic representations of foods from other countries that do not fit the local reality.
- What this study adds: This study brings an adaptation of a graphic representation of foods closer to the local reality, which includes foods that are part of the usual diet of the local population, their recommended portions and a list of substitute foods.
- How this study might affect research, practice or policy This study will contribute to the improvement of food and nutrition education messages and will serve as basic elements in the design of programs, strategies and intervention policies to combat chronic malnutrition that affects the country.

## Introduction

The chronic malnutrition is still a serious public health problem in Mozambique. According to the Household Budget Survey, the prevalence of disease in the country is 38%. ^(1)^

The FAO held an international conference in Rome, in 1992 aiming to identify the strategies and actions to improve food consumption and the nutritional well-being of the population in order to fight for the diseases in the nutrition field. According to this event, the elaboration of food guides for different age groups should be encouraged and, to achieve this objective, each country should plan actions according to its culture and food related health problems. ^(2)^

Food guides are instruments developed by governments or health institutions to guide the population on healthy eating practices. They are based on scientific evidence and adapted to the cultural and nutritional characteristics of each country or region. The main objective of food guides is to promote health and prevent diseases through recommendations on food choice and consumption. ^(3)^

The Food Pyramid is a graphical representation of food guides, a recognizable nutritional tool introduced in 1992. It is shaped like a pyramid to suggest that a person consumes more food from the bottom of the pyramid and less food from the top. ^(4)^

In Mozambique, the graphic representation used for food education is the ‘Poster our food’ drawn by MISAU ^(5)^ which consists of four food groups (Basic Foods, To Protect, To Grow and Concentrated Energy). This work brings a powerful proposal for adapting the food pyramid designed in the United States, in 1992 to the reality of the eating habits of Mozambicans. The Adapted Food Pyramid was built with foods distributed in eight groups (cereals, fruits, vegetables, legumes, milk, meat, fats and sugars) and based on the tables of Mozambican foods compositions, minimum and average caloric consumption of the local population and their eating habits aiming to help nutritionists and other actors of food and nutrition education in Mozambique with improvement approaches and contributing to the reduction of food and nutrition education, chronic malnutrition and other nutritional related diseases that plague the country.

## Methods

After an exhaustive literature review on different graphic representations of foods ^(6)^ and consulting different professionals in the area, the Food pyramid of foods proposed in the United States ^(7)^ was chosen. From this, a new Food Pyramid was built with a description of the groups and minimum and medium portions.

The following stages for the development of the proposal were:

1. Literature review on the different graphic representations of food and choice of elements to adapt and inspire: The review aimed to understand mainly the historical and methodological aspects for the graphic representations designing, experienced specialists in the different areas were also consulted;
2. The identification of Estimates of Minimum and Average Energy Needs of the Population: According to FAO, the minimum energy needs of Mozambicans is 1693 kcal. However, because of not finding the specific average needs for the local population, there was considered the general recommendation of 2,300 kcal with a variation of ± 20%. ^(8-9)^ In this study, the minimum energy requirements was applied to sedentary children and adolescents, while average energy requirements applied to physically active adolescents and sedentary adults. For pregnant women and physically active adults, they should follow specific recommendations.
3. Definition of food groups: The food pyramid was divided into 4 levels, as follows:
  - 1st level: Basic Foods (Derived Cereals and Tubers);
  - 2nd Level: Fruits and Vegetables;
  - 3rd Level: Meat, Milk and pulses;
  - 4th Level: Fats and sugars. Note that the name “Basic Foods” to indicate cereals and derivatives was inspired by the MISAU poster. ^(5)^
4. Definition of portions for each food group: To define the portions, an automated spreadsheet of Microsoft Excel, version 19 was used, which integrates the principles of linear programming analysis of food composition. ^(10)^
5. Designing of food equivalents table: To enable diversification options, a food equivalents table for fixed energy portions was prepared with the help of the spreadsheet and based on the Mozambican composition table. ^(11)^ The Shapiro-Wilk normality test was used to validate the means, assuming the P value greater than or equal to 5% and the standard deviation as a measure of dispersion. Statistical analyses were performed using SPSS, version 27.
6. The standard diet designing: For the validation of the proposal, a standard diet was designed for minimum and average energy value, distributed in four meals (Breakfast, Lunch, Snack and Dinner).

## Results

To check the adequacy of the diet in relation to the recommendation of macronutrient intake, the percentage distribution of macronutrients was calculated for each diet, and for the diet of 1693 kcal, 19% of proteins (10 to 35%), 26% of lipids (20 to 35%) and 55% of carbohydrates (45 to 65%) were obtained. For the 2300 kcal diet, 19% of proteins (10 to 35%), 24% of lipids (20 to 35%), and 57% of carbohydrates (45 to 65%) were obtained, all values are within the recommended range. (12)

**Table 1:** Percentage distribution of macronutrients in diets.

| Diet (Kcal) | Carbohydrates (%) | Lipids (%) | Proteins (%) |
| --- | --- | --- | --- |
| 1693 | 55 | 26 | 19 |
| 2300 | 57 | 24 | 19 |

Source: Automated spreadsheet of Microsoft Excel, version 19.

**Table 2:** 1693 kcal diet.

| FOOD | WEIGHT<br>(g) | HOMEMADE MEASURE | PORTION | GROUP |
| --- | --- | --- | --- | --- |
| **Breakfast** |  |  |  |  |
| Whole milk powder | 51 | 3 Tbsp | 3 | Milk and dairy products |
| Brown sugar | 33 | 1 Tbsp | 1/2. | Sugars |
| Coleslaw | 249 | 18 Tbsp | 1 | Vegetables |
| Canned sardines | 9 | 1 tsp | 1/2. | Meats and eggs |
| Soybean oil | 3 | 1 Teaspoon | 1/2. | Oil and fats |
| Bread | 44 | 1 Ordinary Bread | 1 | Basic food |
| Orange | 134 | 1 Orange the size of a tennis ball | 1 | Fruits |
| **Lunch** |  |  |  |  |
| White rice | 64 | 2 Tbsp | 2 | Basic food |
| Fish | 30 | Hand hand size | 1 | Meats and eggs |
| Butter beans | 29 | 1/2 ladle | 1 | Legumes |
| Raw eggplant | 287 | 8 Tbsp | 1.5 | Vegetables |
| Sunflower oil | 6 | 2 tsp | 1 | Oil and fats |
| Banana | 65 | 1 Unit | 1 | Fruits |
| **Afternoon snack** |  |  |  |  |
| Cookie | 25 | 3 Units | 1 | Basic food |
| Maheu | 46 | 1 small disposable cup | 1/2. | Sugars |
| **Dinner** |  |  |  |  |
| Corn Flour Cup | 32 | 1 Ball* | 1 | Basic food |
| Beef | 17 | 1/2 handle | 1 | Meats and eggs |
| Cabbage | 182 | 1 cup of lunch | 1.5 | Vegetables |
| Sunflower oil | 6 | 1 Teaspoon | 1 | Oil and fats |
| Pineapple | 115 | 1 Slice | 1 | Fruits |

Total Energy = 1659 kcal. Data obtained from Automated spreadsheet of Microsoft Excel, version 19.\* Shell-sized ball, commonly designated locally.
The minimum diet showed the following percentage distribution of energy in the four meals: 33% for breakfast, 36% for lunch, 7% for afternoon snack and 23% for dinner.

**Table 3:**
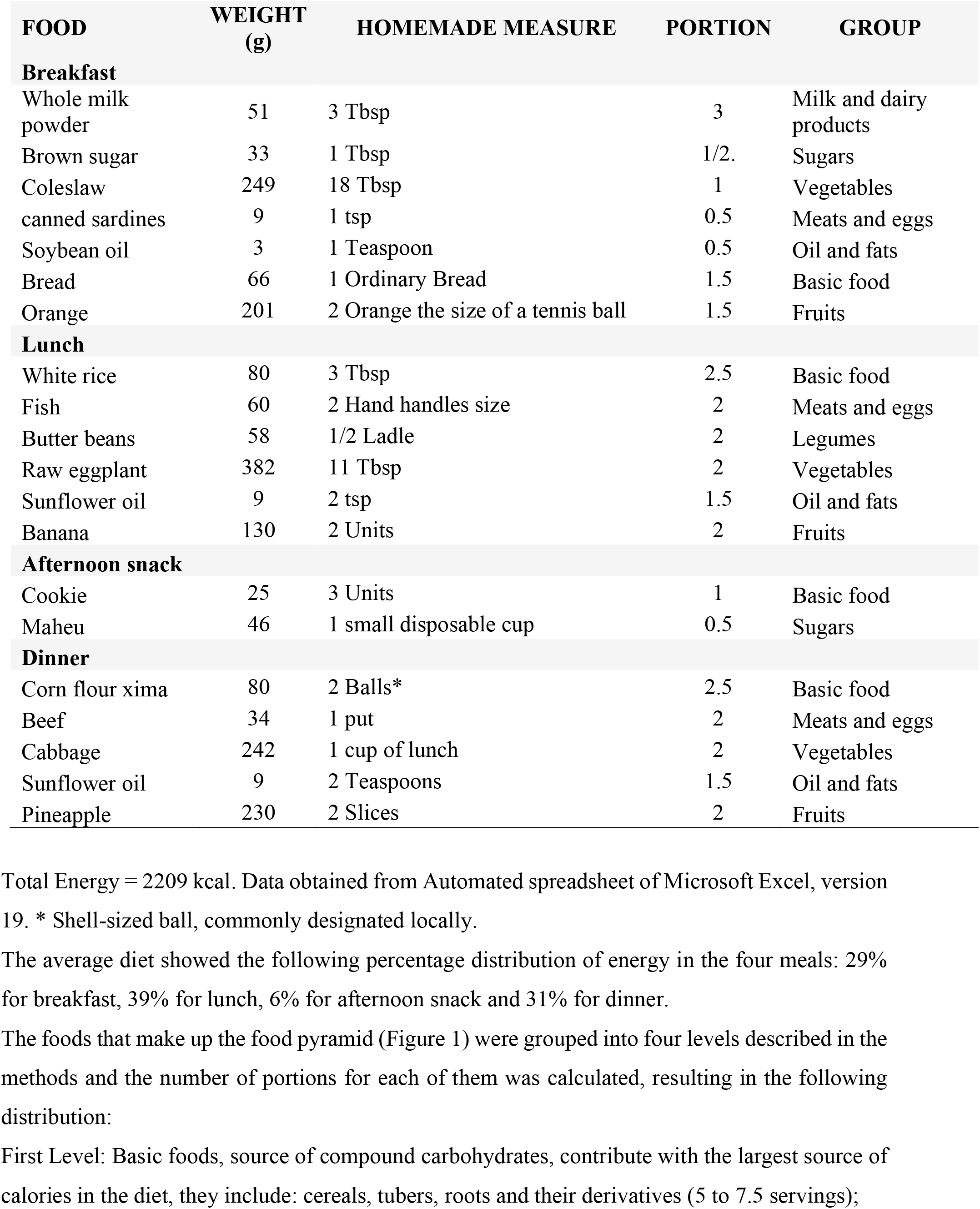
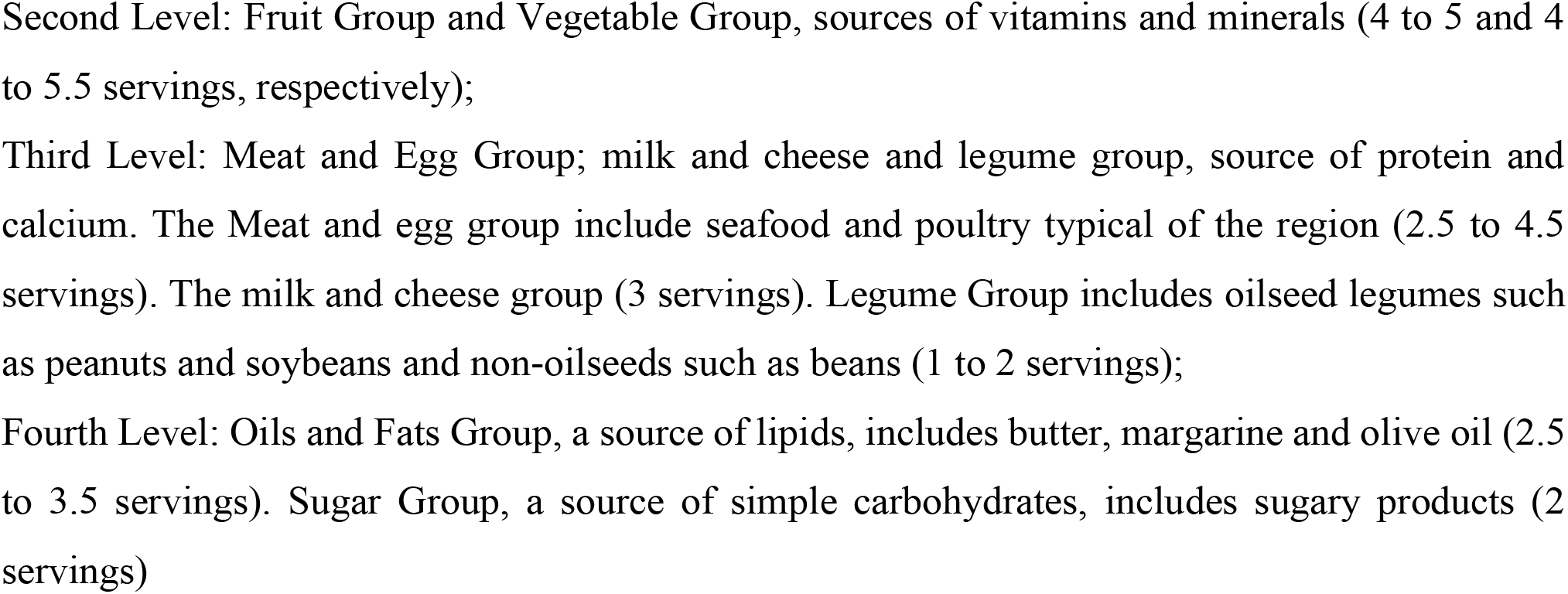
2300 kcal diet.

**Figure 1.**
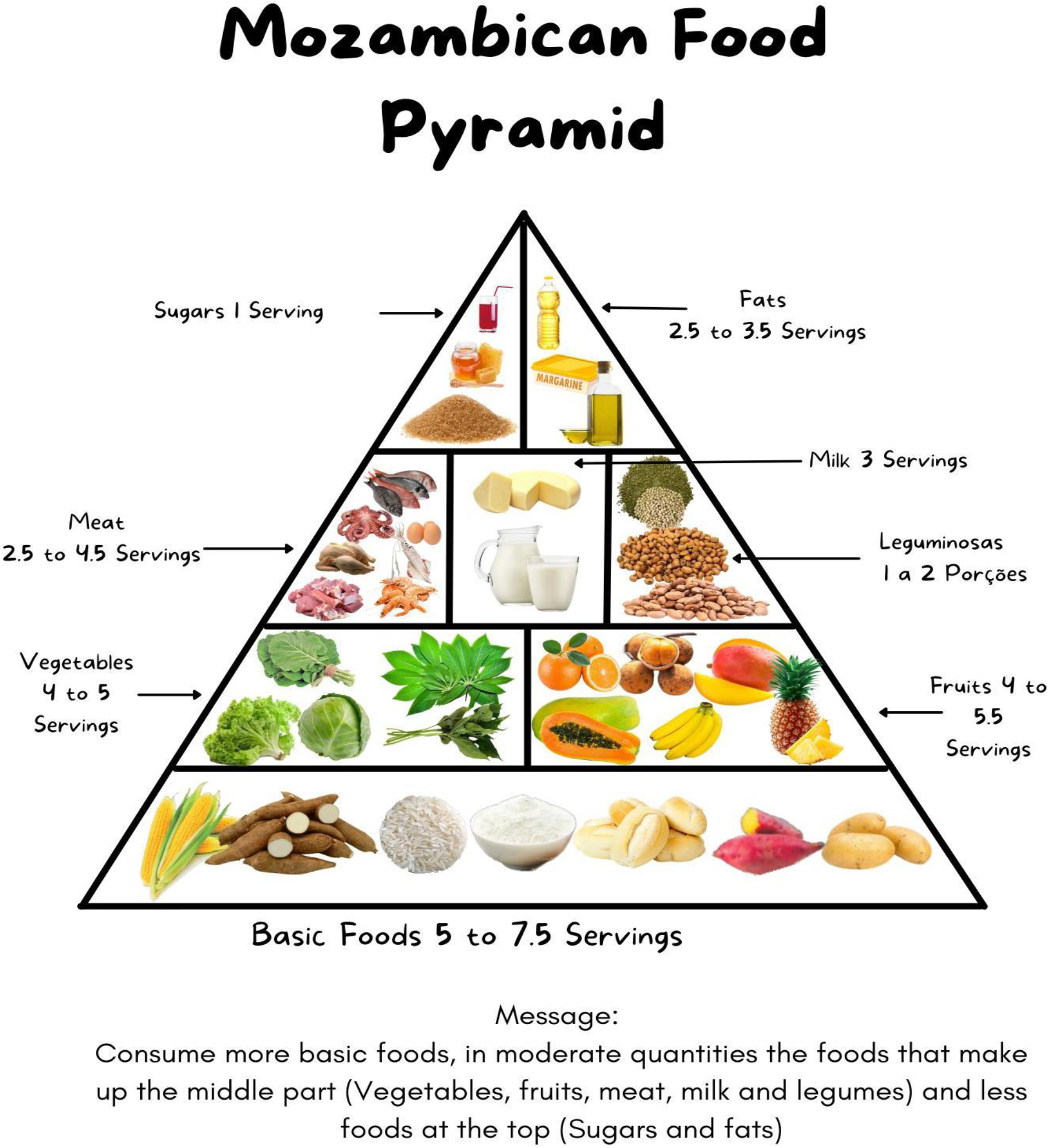
Illustration of the food pyramid drawn. Food pyramid drawn, assembled from photos of local foods, taken from the following sources: PRN^(5)^; Dressup^(13)^; Ifrut^(14)^; Hortfruti^(15)^; Photography^(16)^; Dreamstime^(17)^; Organic^(18)^, Cdifoodservice^(19)^, Istockphoto^(20)^, CozinhaBruta^(21)^, Visãoonoticias^(22)^, Banca13^(23)^, Groupdream^(24)^, CasaBhay^(25)^, Secosdaaldeia^(26)^, Amazon^(27)^, Smilephotoap^(28)^, Ramalho^(29)^, Garden, Herbs^(30)^, Orcapescados^(31)^, Carolina^(32)^, Cienciaviva^(33)^, Sonho^(34)^, Maré^(35)^, Bazara^(36)^, Liudmila^(37)^, Volkov^(38)^, Organic^(39)^, CasaBhay^(40)^, Redemix^(41)^.

## Discussion

This study aimed to design a food pyramid for Mozambique adapted to the local context, based on the old pyramid in the United States, published in 1993 by the Department of Agriculture ^(7)^, as a tool considered in this study as being the one that comes close to the local reality.

The study conducted by Fernandes et al. ^(42)^ aimed to design a food pyramid that represented all the countries of Sub-Saharan Africa. However, due to the diversity of cultures and the lack of information on defined portions, its application in the local context as an integral part of it may be a challenge. Because of that, and with little information in the literature on the specific food pyramid for Mozambique, this study aimed to close the gap by bringing a proposal for this tool. At the local level, there is a poster designed by the Ministry of Health ^(5)^, but the similarity of the graphic representation proposed by Fernandes ^(42)^ does not provide information about the portions of food, thus subjectivizing the messages of food education. The size of the portions defined in this study took into account the eating habits described by Machamba ^(43)^, food availability and the diversity of crops in which Mozambique is rich ^(44)^.

The adapted pyramid was designed with diets of minimum and average caloric needs while the original one was designed with three values (minimum energy needs, 1600 kcal. average, 2200 kcal. and maximum, 2800 kcal). The group of cereals and derivatives constitutes the basis of the diet of Mozambicans, which, according to Machamba ^(43)^, with an average consumption of 725 g/day, followed by tubers, with an intake of 331, in this study, the consumption of cereals in portions is about 240 g for cereals and 1088 g for tubers, because in the usual quantities the contribution of carbohydrates exceeds the recommended standards. For the fruit and vegetable group, we tried to approximate the portions defined in this study (4 to 5 portions) with those recommended by the WHO ^(45)^ (consumption greater than or equal to 400 g/day). The Meat group is the least consumed group in Mozambique, once, according to Machamba ^(43)^, the average daily consumption is only 11 g, a fact that may be contributing to the levels of chronic malnutrition that plagues the country, especially in children under 5 years, as revealed by INE data ^(46)^. To fill this need, this study defined the consumption of 4 to 4.5 daily servings of this food group. The group of milk and dairy products deserved special attention because it is an important source of calcium and, according to the WHO ^(18)^, is associated with the prevention of bone diseases and cancer. With the consumption of the portions recommended in the pyramid of this study (3 daily servings), it is possible to supply 453 mg to 528 mg of this mineral, which corresponds to 50% of the Recommended Dietary Allowance ^(19)^. The legume group in the United States pyramid was included in the meat group, but given the importance of the group in the local diet, which serves as an alternative source of protein, especially in populations living in rural areas of Mozambique, due to its low production cost and affordability ^(20)^, it was placed separately as a separate group, with a recommendation of between 1 and 2 daily portions. The group of sugars, oils and fats constituted the group at the top of the pyramid that should be consumed with caution. For the Sugars group, consumption is limited to less than 10% of total calories because its excessive consumption is associated with chronic non-communicable diseases. ^(18)^ In this study, the caloric contribution of this group is approximately 1%. Similarly to the group of Fats and Oils, consumption is limited to between 15 and 30% of the total value of energy ^(18)^, in this study, the diets of this group contribute with 19% of the total calories of the diets.

## Conclusion

The food pyramid developed is presented as a guidance tool for nutritionists and other actors in food and nutrition education in Mozambique. The instrument was adapted considering factors such as availability, reference dietary intake, and eating habits of the local population. Attention was paid to energy balance when developing standard diets with minimum calories consumed by the general population and those recommended on average. The portion sizes and the list of food equivalents are adapted to the population’s eating habits and frequency of food consumption. Special attention was given to the milk group, the portions and their size were defined to supply 50% of the dietary reference intake of calcium. Further studies are needed to validate and implement the instrument so that it becomes an effective and practical tool.

## Data Availability

All data produced in the present study are available upon reasonable request to the authors

## Appendix

## Appendix 1

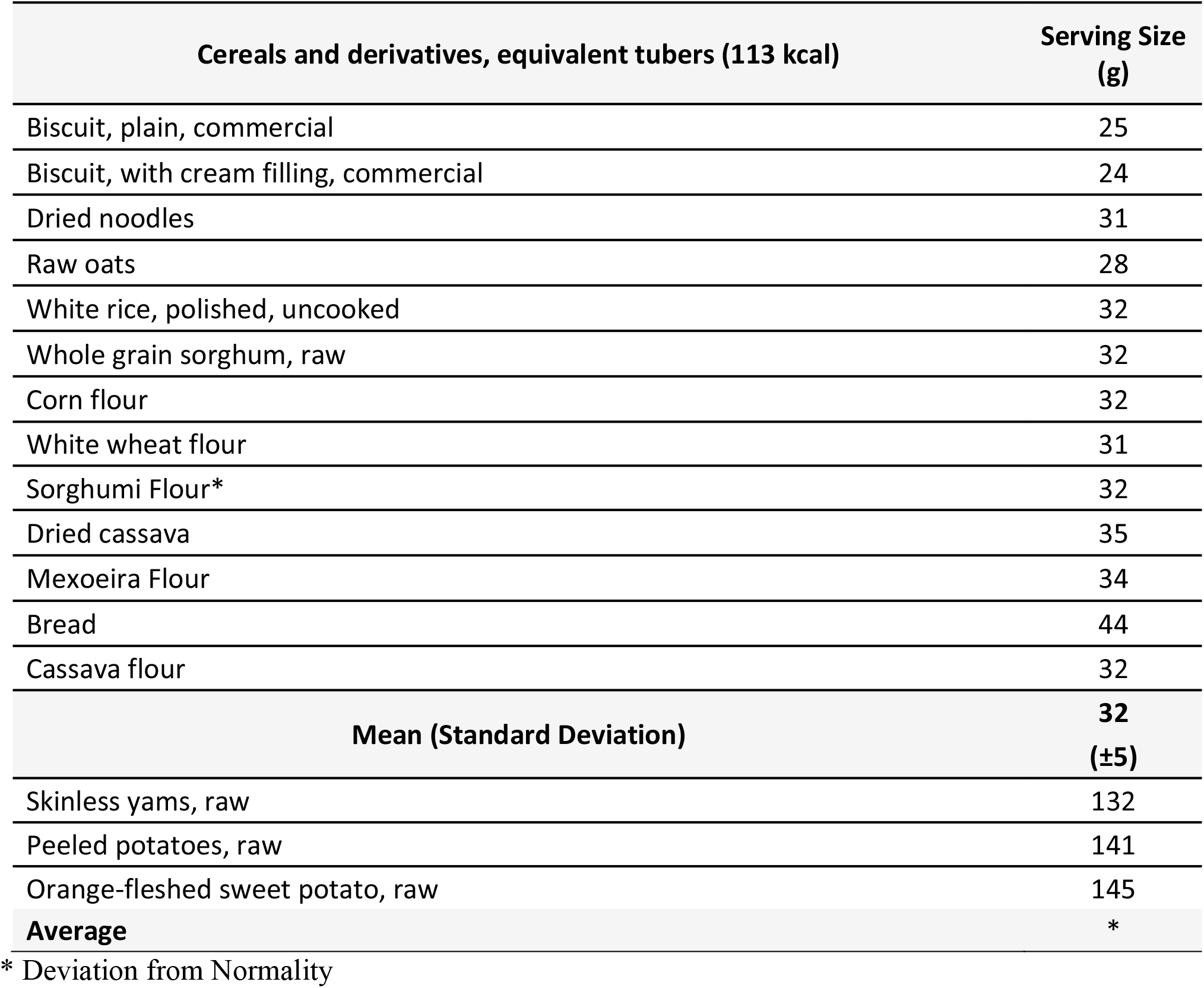

## Appendix 2

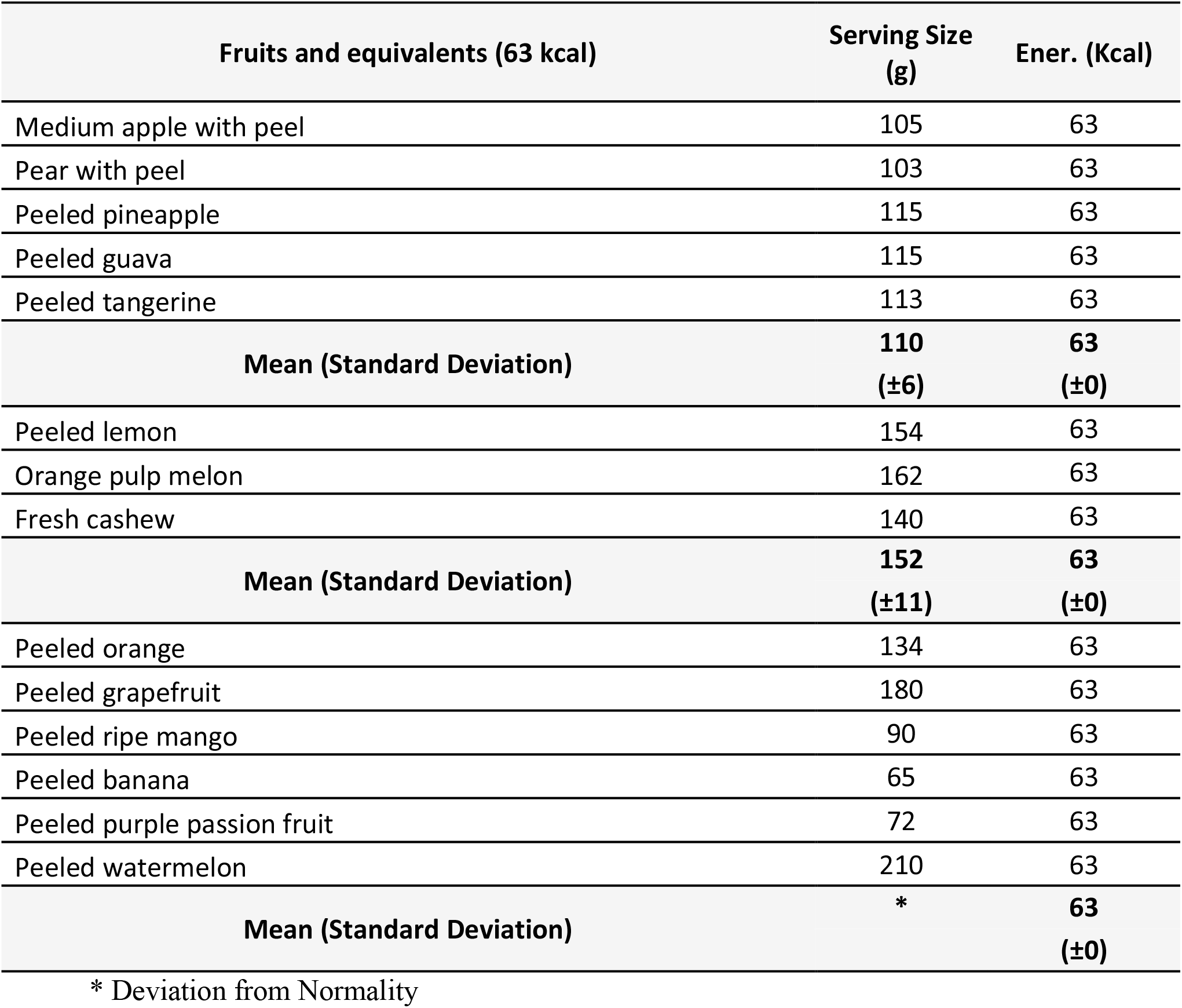

## Appendix 3

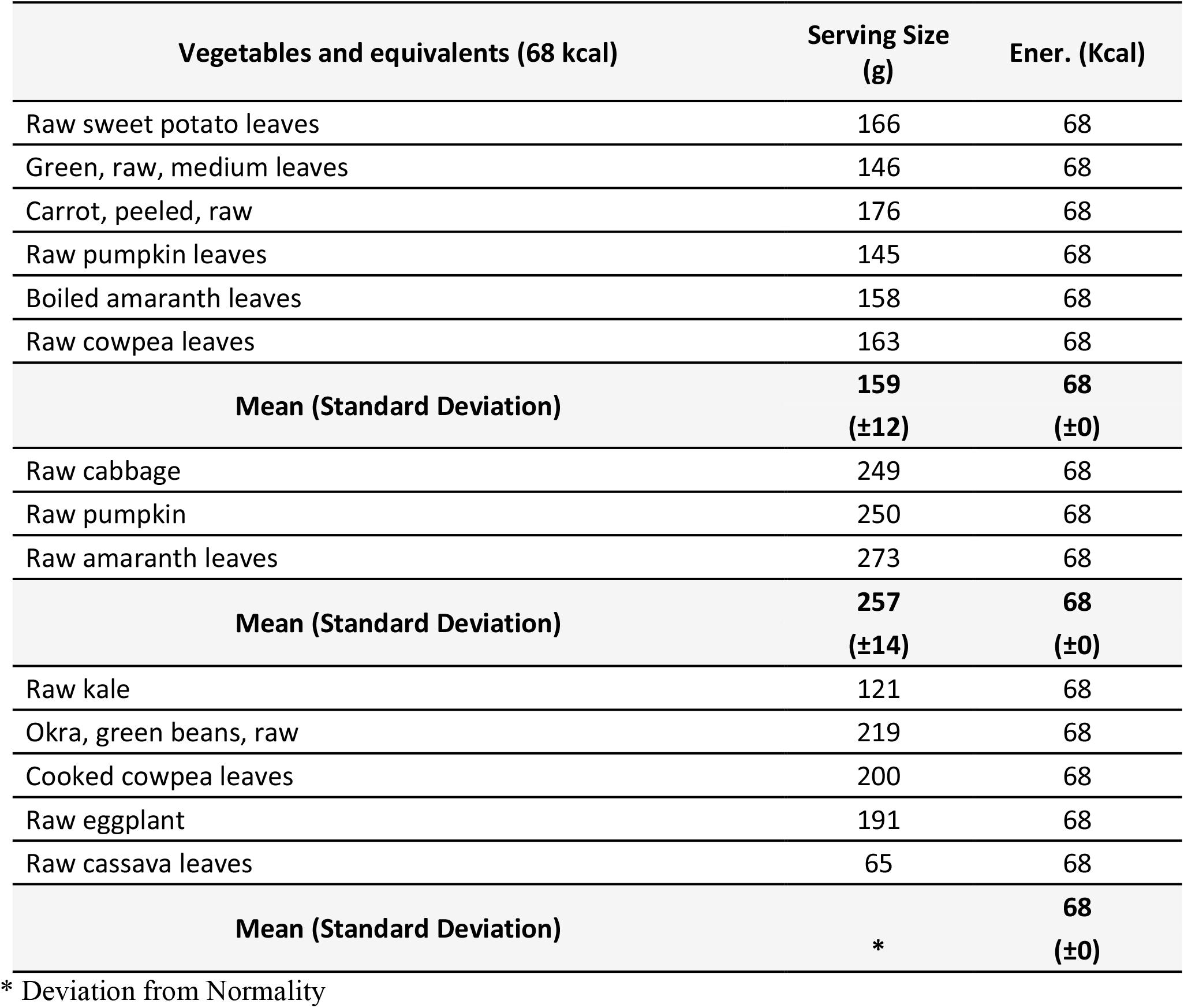

## Appendix 4

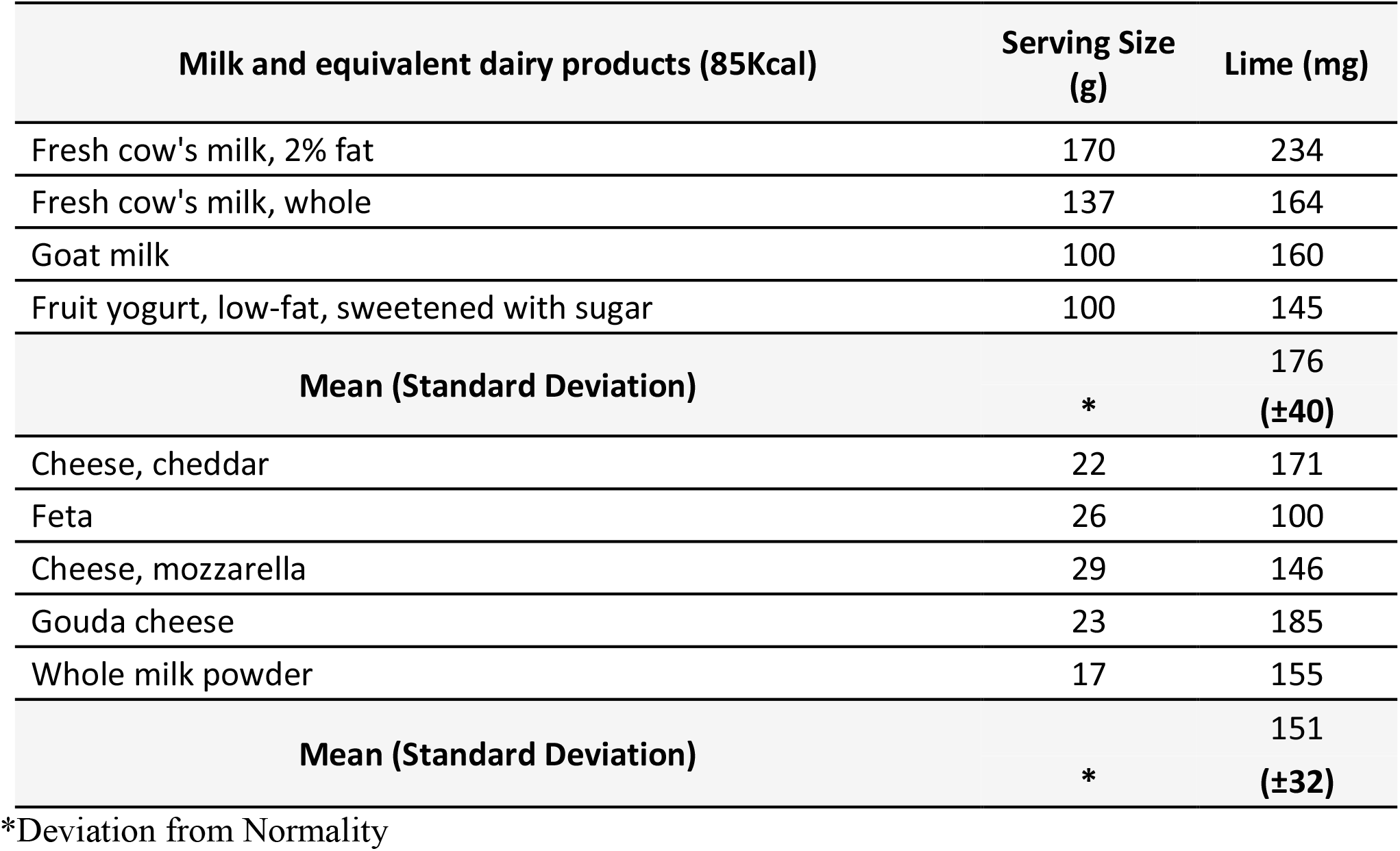

## Appendix 5

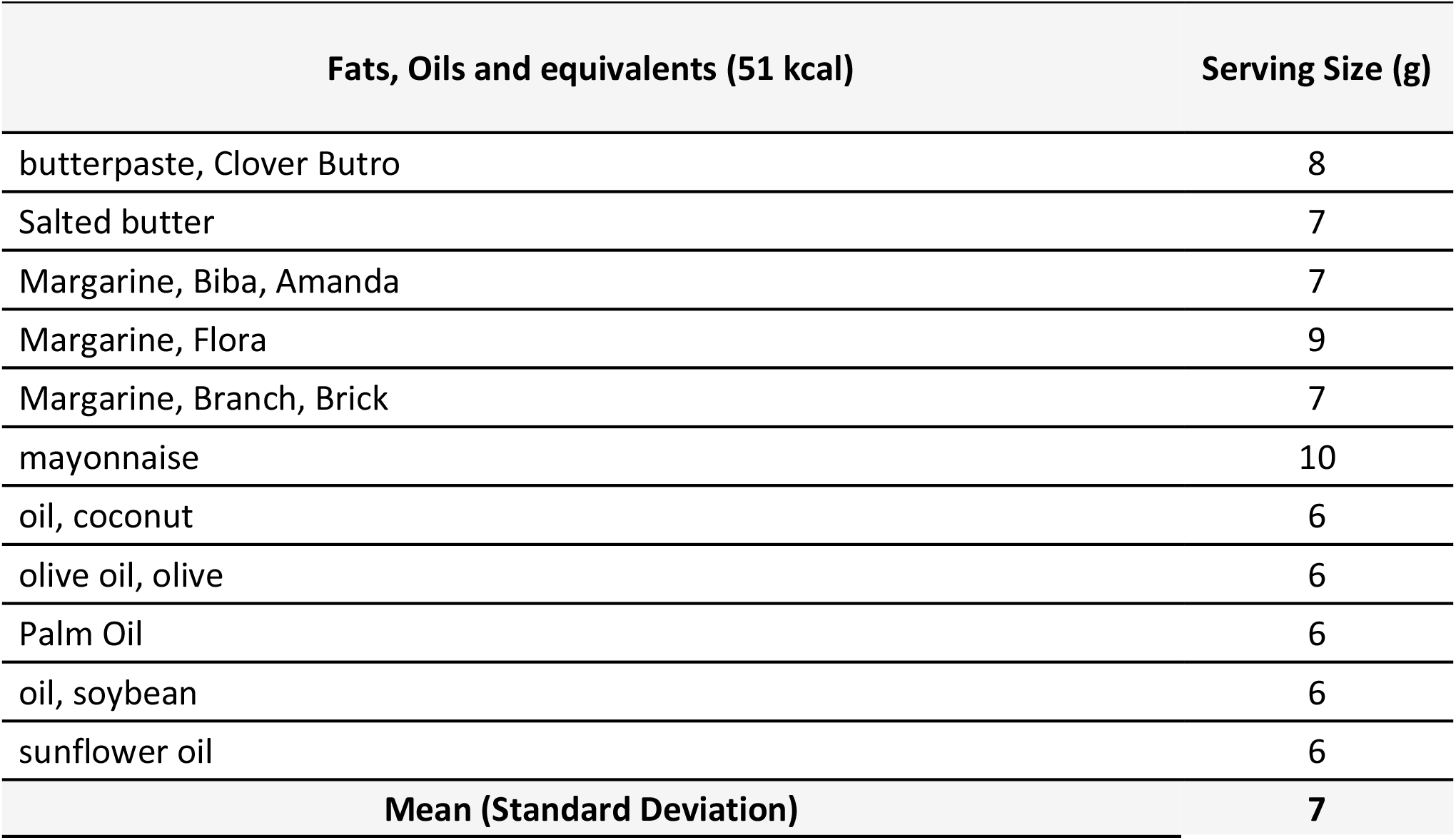
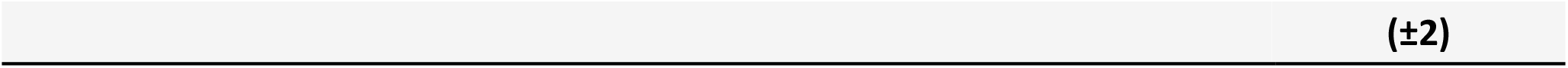

## Appendix 6

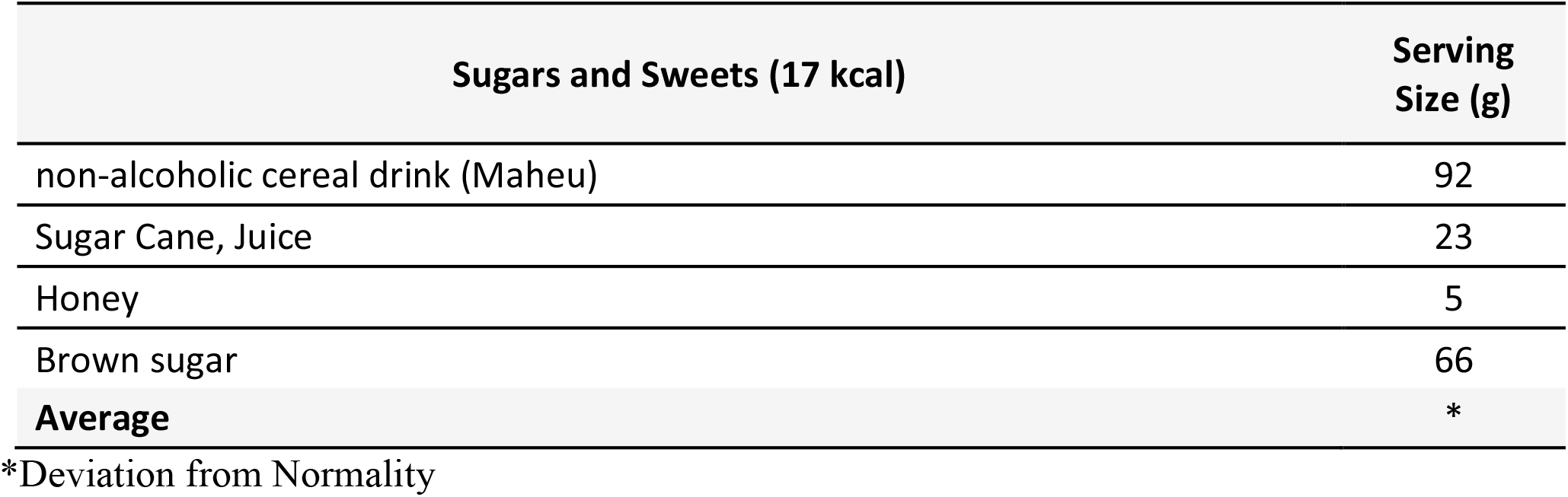

